# Genetics of Unexplained Sudden Cardiac Death in Adult Caucasian and African American Individuals Living in the State of Maryland

**DOI:** 10.1101/19007344

**Authors:** Liang Guo, Sho Torii, Raquel Fernandez, Ryan E. Braumann, Ka Hyun Paek, Daniela T. Fuller, Christina M. Mayhew, Roya Zarpak, Hiroyuki Jinnouchi, Atsushi Sakamoto, Yu Sato, Hiroyoshi Mori, Matthew D. Kutyna, Parker J. Lee, Leah M. Weinstein, Carlos J. Collado-Rivera, Neel V. Gadhoke, Bakr B. Ali, Dheeraj R. Atmakuri, Roma Dhingra, Emma LB. Finn, Mack W. Bell, Anne Cornelissen, Salome H. Kuntz, Joohyung Park, Robert Kutys, Libin Wang, Susie N. Hong, Anuj Gupta, Frank D. Kolodgie, Maria E. Romero, Braxton D. Mitchell, Dipti Surve, David R. Fowler, Charles C. Hong, Renu Virmani, Aloke V. Finn

**Affiliations:** CVPath Institute, Gaithersburg, Maryland, USA; University of Maryland School of Medicine, Department of Cardiovascular Medicine, Baltimore, Maryland, USA; Office of the Chief Medical Examiner, Baltimore, Maryland, USA

## Abstract

**Background:** Unexplained-sudden cardiac death (SCD) describes SCD with no cause identified after a comprehensive autopsy and toxicologic examination. Genetic testing helps to diagnose inherited cardiac diseases in unexplained-SCD, however, the relationship between pathogenic or likely pathogenic (P/LP) variants of inherited cardiomyopathies and primary electrical disorders (PED) and risk of unexplained-SCD in adults living the United States has never been systematically examined.

**Methods:** We performed sequencing of 29 cardiomyopathy and 39 arrhythmia genes in 413 African-Americans and Caucasians (≥18 years-old) who died of unexplained-SCD (median age; 41 years-old, 37% females, 50% African-Americans) and whose hearts were found to have no gross pathological finding upon referral to our institution for pathologic consultation from the State of Maryland Medical Examiner. We examined P/LP variants in these genes to study the association between presence of these variants and unexplained-SCD.

**Results:** 143/413 (34.6%) subjects had variants considered P/LP for cardiomyopathy and/or PED (i.e. Brugada Syndrome (BrS), long QT syndrome (LQTS), and arrhythmogenic right ventricular dysplasia (ARVD)). In total, 102 (24.7%) subjects harbored 86 P/LP variants for cardiomyopathies and 60 (14.5%) subjects carried 76 P/LP variants for PED. The following pathogenic variants were identified: 68 P/LP variants for hypertrophic cardiomyopathy (HCM) in 79/413 (19.1%) subjects, 18 P/LP variants for dilated cardiomyopathy (DCM) in 22/413 subjects (5.3%), and 76 P/LP variants in 60/413 (14.5%) for PED. There were greater than 121.0- and 138.5-fold median enrichments (431.4- and 200.0-fold cumulative enrichments) in these cardiomyopathy and arrhythmia variants in victims of unexplained SCD versus the general population, respectively. Among these P/LP positive carriers, combinations of conditions were found, including 14/413 (2.4%) having both HCM and PED variants, and 5/413 (1.2%) with DCM and PED variants. African Americans (AA) and Caucasians were equally likely to harbor P/LP variants (32.7% versus 36.6%, p=0.5), but AA had a higher frequent variants of unknown significance.

**Conclusions:** This study represents the largest examination reported on the association between cardiomyopathy and arrhythmia P/LP genetic variants and unexplained-SCD in adults with no gross abnormality on rigorous pathological examination. Nearly one-third of those with unexplained-SCD were carriers of P/LP variants. Our findings with respect to both the association of unexplained SCD with cardiomyopathy genes and race-specific genetic variants suggest new avenues of study for this poorly understood entity.

## INTRODUCTION

The incidence of sudden cardiac death (SCD) in the United States (US) ranges from 180,000 to 450,000 cases annually ^1, 2^. Coronary heart disease (CHD) is the most common substrate underlying SCD in the western world, being responsible for 50-75% of SCDs. While autopsy studies may lead to a diagnosis of structural heart disease in many of the remaining cases (e.g. SCD subjects without CHD), in 30-40% (so-called unexplained-SCD) the cause still remains uncertain despite toxicologic and histopathologic analysis ^2, 3^.

Molecular autopsy improves diagnostic accuracy in subjects with normal cardiac autopsy. In the young, primary electrical disorders (PED) are a well-recognized cause of SCD in the absence of structural heart disease. Although several cardiomyopathy gene variants have also been reported to increase risk for SCD; whether these variants can accurately predict future SCD risk, especially in those without overt structural heart disease, remains controversial ^4, 5^. Recently European recommendations were published integrating genetic testing into multidisciplinary management of SCD. These guidelines emphasized the importance of genetic testing and appropriate information provision of affected individuals, as well as their relatives ^6^.

Most studies of unexplained SCD conducted to date have concentrated on small numbers of young subjects (i.e. <35 years of age) from Australia, New Zealand, Denmark, Netherlands, Spain, United Kingdom, France, and South Korea ^7-13^, all relatively homogenous populations without the racial diversity seen in the US. Even though the incidence of sudden death increases dramatically with age and is higher in men than women, our understanding of the causes of unexplained SCD in adults, especially those of African Americans descent, is limited ^14^.

Here we conducted the largest and most systematic genetic examination of adult African American and Caucasian individuals in the US who died and were referred for autopsy with suspected SCD but had normal cardiac pathologic findings. Our goal was to understand the association between pathogenic variants for PED and cardiomyopathies and risk of unexplained-SCD in adult individuals living in the US.

## METHODS

### Study Design and Oversight

Between 1995 to 2015, CVPath Institute was referred a total of 5,262 hearts from cases of unexpected sudden death from the Office of the Chief Medical Examiner of the State of Maryland (OCME-MD). (Unexpected sudden death cases in the state of Maryland are routinely referred to our Institute for consultation at the discretion of the medical examiner). At OCME-MD, a complete and comprehensive autopsy and toxicologic analysis is performed in all referred decedents up to 50 years old and in cases over 50 years old without evidence of possible drug/alcohol abuse. For every case, the heart was systematically evaluated with detailed histopathological analysis performed by an experienced cardiac pathologist at CVPath. To examine race-based differences, only African American or Caucasian individuals over 18 years old were analyzed in the current study (n=4,270). The racial origin of subjects was identified from the OCME-MD report through inquiry of family members. Subjects with clear non-cardiac cause of death as well as cases with lack of detailed information were excluded from the study (n=950). Subjects with SCD due to sudden coronary death (n=1,875; i.e. acute coronary syndrome, ≥75% stenosis in any major epicardial coronary artery, or previous bypass or stent placement) and other known causes of sudden cardiac death (n=762; i.e. cardiomyopathy (e.g. hypertrophic, dilated or arrhythmogenic right ventricular cardiomyopathies, etc.), significant valvular disease, congenital heart disease, myo/pericarditis, or infective endocarditis) were excluded from the study. See Figure 1 for study flowchart.

**Figure 1.**
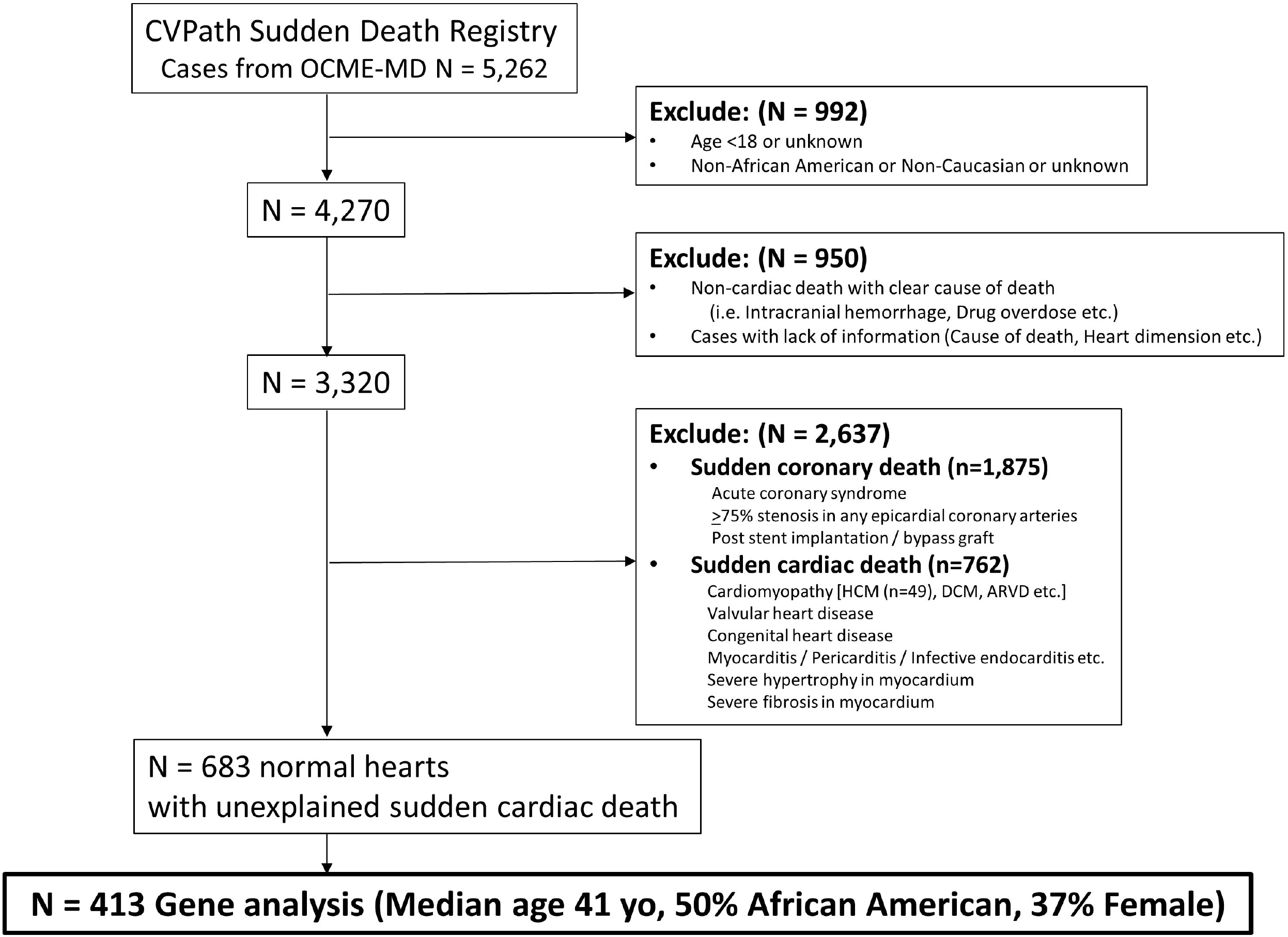
Study Flow Diagram of the Investigation of Genetics of cardiomyopathy and arrhythmia genes in Subjects with Unexplained Sudden Cardiac Death. From 5,262 cases of suspected sudden cardiac death collected from 1995-2015 from the Office of the Chief Medical Examiner of the State of Maryland (OCME-MD), a total of 683 had normal cardiac autopsy were classified as unexplained sudden cardiac death. From these 413 cases were chosen for DNA extraction from each racial group (i.e. African American and Caucasian) to match the racial makeup of the original 683 subjects. (See Methods for more study design details) HCM= hypertrophic cardiomyopathy, DCM= Dilated cardiomyopathy, ARVC= Arrhythmogenic right ventricular cardiomyopathy

### Assessment of Hearts to rule out any known cause of death

More details of the histopathologic analysis are given in the Methods section in the Supplemental Materials. All specimens had been fixed in 10% buffered formalin. The epicardial coronary arteries were sectioned at 3 to 4 intervals to rule out any significant atherosclerosis. The base of the heart at the level of the tip of the mitral valve was used to measure the left ventricle (LV) free wall, ventricular septal thickness, and right ventricular wall thickness followed by left ventricular cavity diameter excluding the pectinate and papillary muscles. In total 6 sections of myocardium (anterior, posterior, and lateral LV, ventricular septum, anterior and posterior wall of the right ventricle) were routinely taken transversely, embedded in paraffin, and stained with hematoxylin and eosin (H&E) stain for histologic evaluation. Histologic examination was performed to rule out any infiltrative process or any myofiber disarray of the myocardium, intramyocardial small vessel disease of interstitial or focal fibrosis. The presence of any of these cardiomyopathic processes resulted in exclusion from the normal heart category. Wall thickness measurements from subjects with normal hearts were consistent with previous autopsy studies of “normal hearts” ^15^. (Autopsy wall thicknesses overall are thicker than that reported by echocardiographic measurement in living patients which is consistent with prior studies and values from previous autopsy studies of normal hearts ^16^.)

### Definition of Normal Hearts

“Normal” heart was defined as an individual dying of natural causes with no evidence of heart disease following a complete gross and histopathological evaluation. Assessment of the myocardial sections ruled out the presence of any cardiomyopathic process. In addition, microscopic examination also helped to rule out severe myocyte hypertrophy (defined as cardiac myocyte thickness greater than 25 µm ^17^), and severe fibrosis in the myocardium (area of fibrosis >3% of the section) (Figure 1). Suspected unexplained-SCD was defined as symptoms commencing within 1 hour of death (with or without witnessed arrest) or death occurring within 24 hours after the victim was last seen alive in their normal state of health, and in whom a clear cause of death (CoD) could not be established after a complete and comprehensive autopsy examination (including cardiac examination). All cases of SCD with normal autopsy were adjudicated to be unexplained by three cardiologists (ST, CCH, AVF) and a cardiac pathologist (RV). The protocol for the study was approved by the Institutional Review Board of the CVPath Institute.

### DNA Collection, Next-generation Sequencing and Data Analysis

Genomic DNAs were extracted from either fresh frozen tissue and FFPE tissue blocks using DNeasy Tissue kit and QIAamp DNA FFPE Tissue Kit (Qiagen), respectively. TruSeq Custom Amplicon Low Input was used to construct the libraries using custom design cardiomyopathy gene panel (Illumina, San Diego, CA). The libraries were multiplexed and sequenced on HiSeq 550 Rapid Run mode (Illumina). Fastq sequencing data were analyzed using TruSeq Amplicon workflow Version 3.0.0 on BaseSpace platform (Illumina), including Isas (analysis software) Version 1.1.7.9.271 + TSAv3, SAMtools Version 1.2, Isas Smith –Waterman-Gotoh (Aligner) Version 6.2.1.25+ develop, Pisces Variant Caller Version 5.2.1.22, and Illumina Annotation Engine 1.5.3.82 against human genome GRCh37/hg19. Alignment was performed using the banded Smith-Waterman algorithm in the targeted gene regions. Primary Electrical Disorders (PED) MASTR Plus was used to construct the libraries for arrhythmia gene panel (Muiltiplicom, Agilent, Belgium). The libraries were multiplexed and sequenced on HiSeq 550 Rapid Run mode (Illumina). Fastq sequencing data were analyzed using Sentieon Workflow pipeline on BaseSpace platform (Illumina), which uses the same mathematics used in the Broad Institute’s BWA-GATK HaplotypeCaller Best Practice Workflow pipelines. The genomic VCF files and BED files from both cardiomyopathy and PED panels were used for genomic variants callings analyses on SVS software Version 8.0 (Golden Helix, Bozeman, MT) and Integrative Genomics Viewer Version 2.4 (IGV) (Broad Institute).

### Variant filtering and classification and comparison with general population

A list of the 29 target cardiomyopathy genes and 39 target arrhythmia genes are shown in Supplemental Table 1. P/LP variants with a minor allele frequency (MAF) of greater than 0.01% in the Exome Aggregation Consortium (ExAC), TOPMED, and GnomAD were excluded from the analysis. The ClinVar database (July 2019 release, NIH) was used to determine P/LP variants, and all the P/LP variants were individually checked for the accuracy of classification. All the P/LP variants with a read depth <15 were validated using Taqman genotyping assays (Thermo Fisher, Waltham, MA). After variant classification, MAF of each benign, variants of unknown significance (VUS), P/LP variants was compared with that of the general population using allele frequency data from TOPMED, ExAC, GnomAD, 1000G, and UK10K.

### Detailed evaluation of myocardial fibrosis

All the sections of myocardium in left ventricle (septal, anterior, posterior, lateral wall) in consecutive cases whose myocardial tissues were available, and samples with or without P/LP in HCM gene variants were stained with Masson Trichrome stain to evaluate for myocardial fibrosis as described in the results section. Whole areas with myocardial fibrosis (which stains blue with Masson Trichrome stain) were measured using ZEN software (Zeiss, Oberkochen, Germany). See Methods section in the Supplemental Materials for more details of the evaluation of myocardial fibrosis.

### Statistical analysis

Results for continuous variables with normal distribution were expressed as mean ± SD. Normality of distribution was tested by the Shapiro–Wilk test. Variables with non-normal distribution were expressed as median and 25th to 75th percentiles. Categorical data were expressed as numbers (%) and analyzed by Chi-square test or Fisher’s Exact tests. Student t test was used to analyze the significance of differences for continuous variables with normal distribution, whereas comparisons of variables with non-parametric distribution were performed by the Kruskal-Wallis test. A p value of <0.05 was considered statistically significant. JMP software version 13.0 (SAS, Cary, NC, USA), SPSS software version 22 (IBM, Armonk, NY, USA), and GraphPad Prism version 8.0 (GraphPad Software, San Diego, CA, USA) were used for statistical analyses.

## RESULTS

### Demographics and Heart Measurements of the Study Cohort

The study design is shown in Figure 1. From 1995-2015 the CVPath Sudden Death Registry collected 5,262 autopsy hearts from OCME-MD. A total of 992 subjects were excluded because they did not meet study demographic criteria (e.g. Age<18, non-African Americans or non-Caucasians). Another 950 subjects were excluded because the CoD was a non-cardiac death, or there was a lack of detailed information about final diagnosis or heart dimensions. Another 2,637 subjects were excluded because the CoD was a known cardiac cause such as obstructive CAD (including but not limited to acute coronary syndromes), valvular disease, cardiomyopathy, myocarditis, etc. Overall, 683 subjects with “normal” cardiac autopsy were adjudicated to have suffered unexplained-SCD. Among these, 413 cases with acceptable DNA quality for sequencing were chosen to approximately match the racial makeup of the original 683 subjects. The demographic and clinical characteristics of 413 patients are shown in Supplemental Table 2. A total of 63% of the patients were male, and the mean age at death was 41 (29-48) with 208 being African Americans and 205 Caucasians.

### Frequency of P/LP Genetic Variants for Cardiomyopathy and PED Disorders in the Study Cohort

413 subjects with suspected unexplained-SCD were analyzed for 29 cardiomyopathy and 39 primary arrhythmia gene variants (Supplemental Table 1). In total, 143/413 (34.6%) subjects had variants considered P/LP for cardiomyopathy (HCM and DCM) and/or PED (i.e. Brugada Syndrome (BrS), LQTS, and ARVD). 102 (24.7%) subjects harbored 86 P/LP variants for cardiomyopathies and 60 (14.5%) subjects carried 76 P/LP variants for PED. It is also worth noting that 19 (4.6%) subjects carried variants for both conditions (Figure 2A-B). P/LP variants in MYH7 (n=28/86, 33.3%) and MYBPC3 (n=28/86, 33.3%) were the most frequent cardiomyopathy genes (Figure 2C), while variants in SCN5A (n=18/76, 23.7%), KCNQ1 (n=14/76, 18.4%), KCNH2 (n=12/76, 15.8%), DSP (n=9/76, 11.8%), and RYR2 (n=8/76, 10.5%) were the most frequently found arrhythmia genes consistent with BrS, LQTS, or ARVD (Figure 2D). More than 90% of P/LP variants in MYH7, TNNT2, TNNI3, SCN5A, KCNQ1, and KCNH2 were missense variants, on the other hand, 33/37 (89%) P/LP variants in MYBPC3 and DSP were truncating variants, including stop gained and different splice variants (Supplemental Figure 1), which is consistent with the previous reports describing the type of mutations seen in these disorders ^18, 19^. The clinical characteristics, circumstances of death, CoD, and relevant genetic information for every subject with P/LP variants are provided in Supplemental Table 3.

**Figure 2.**
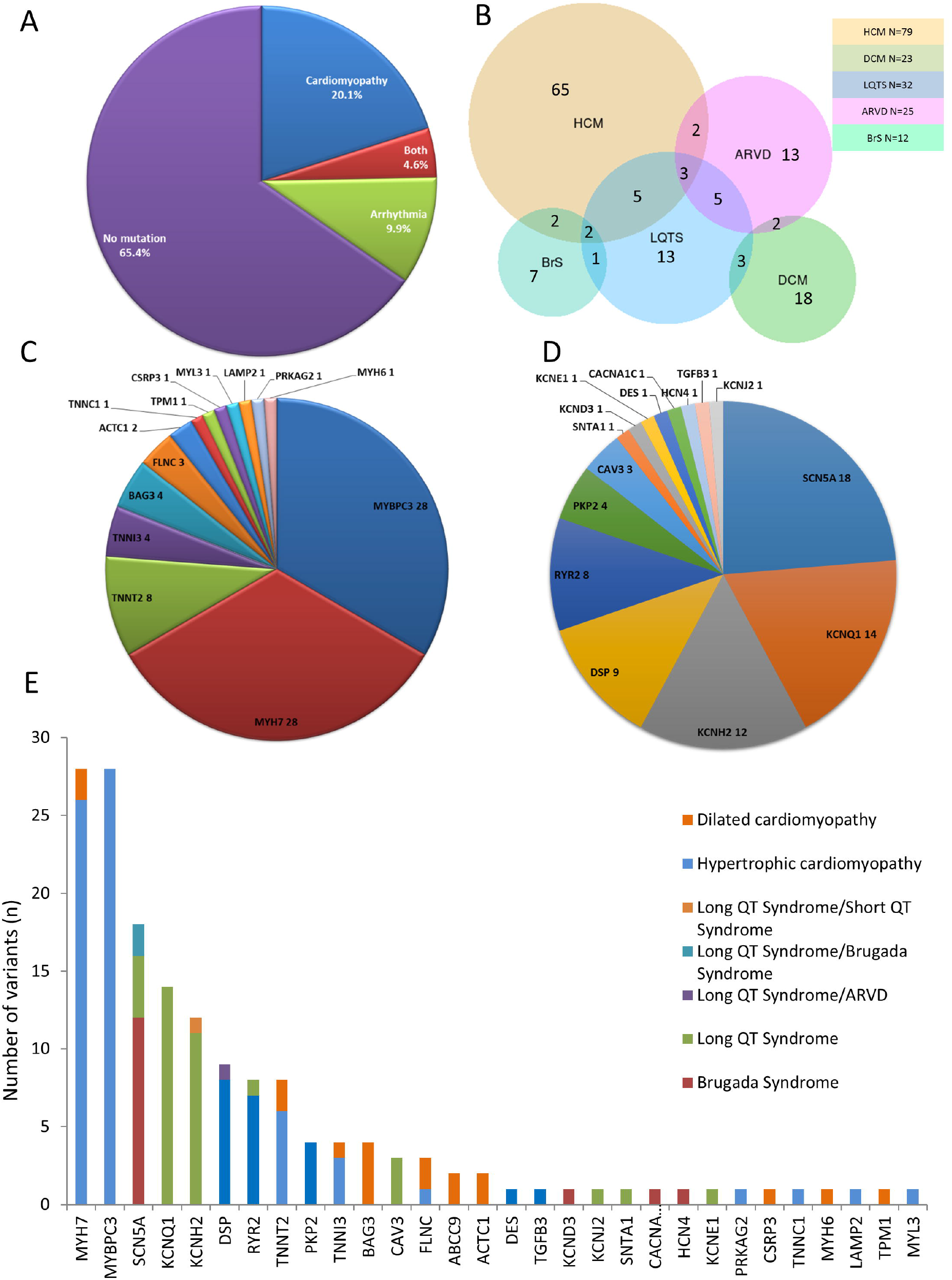
Results of Genetic Testing of cardiomyopathy and arrhythmia genes in Subjects with Unexplained Sudden Cardiac Death. Among 413 subjects analyzed for cardiomyopathy and arrhythmia gene variants, 86 variants for cardiomyopathy and 76 variants for PED considered pathogenic or likely pathogenic (P/LP) were found from 143 subjects. **A**. 102/413 and 60/413 carried P/LP variants for cardiomyopathy and arrhythmia, respectively. Among them, 19/413 subjects carried combination of variants in both conditions. **B**. Each condition and associated P/LP variant carriers are shown in the Venn diagram. **C**. Pie chart illustrating overview of genes harboring variants for cardiomyopathy. More than two-third of P/LP variants in MYBPC3 and MYH7, **D**, variants in SCN5A, KCNQ1, KCNH2, DSP, and RYR2 are the most frequent found P/LP variants for BrS, LQTS, ARVD. **E**. Genes found with P/LP variants and associated conditions are shown.

When the MAF for benign and P/LP variants within genes examined in the current study were compared to the general population, MAF for benign variants showed a linear relationship between our SCD cohort and the general population (R^2^=0.8618, p<0.0001) (Figure 3A). In contrast, MAF for P/LP was non-linear between SCD study cohort and general population (R^2^=0.002, p=0.9851), and was highly enriched in the study cohort (Figure 3B). The median fold enrichment in our SCD cohort for P/LP variants compared with general population were 121.0 and 138.5 [interquartile range (IQR) 63.0 - 272.0 and 63.3 – 297.9] for cardiomyopathy and arrhythmia P/LP variants, respectively, which were significantly greater than for benign variants, which had a median fold enrichment of 1.2 (0.7-1.8, p=ns)) (Figure 3C). Cumulatively, there were 431.4- and 200.0-fold enrichment in cardiomyopathy and arrhythmia variants in victims of unexplained SCD, respectively (cumulative MAF of 0.1842 and 0.1436 in unexplained SCD cohort versus cumulative MAF of 0.000427 and 0.000718 in the general population for cardiomyopathy and arrhythmia P/LP variants, respectively).

**Figure 3.**
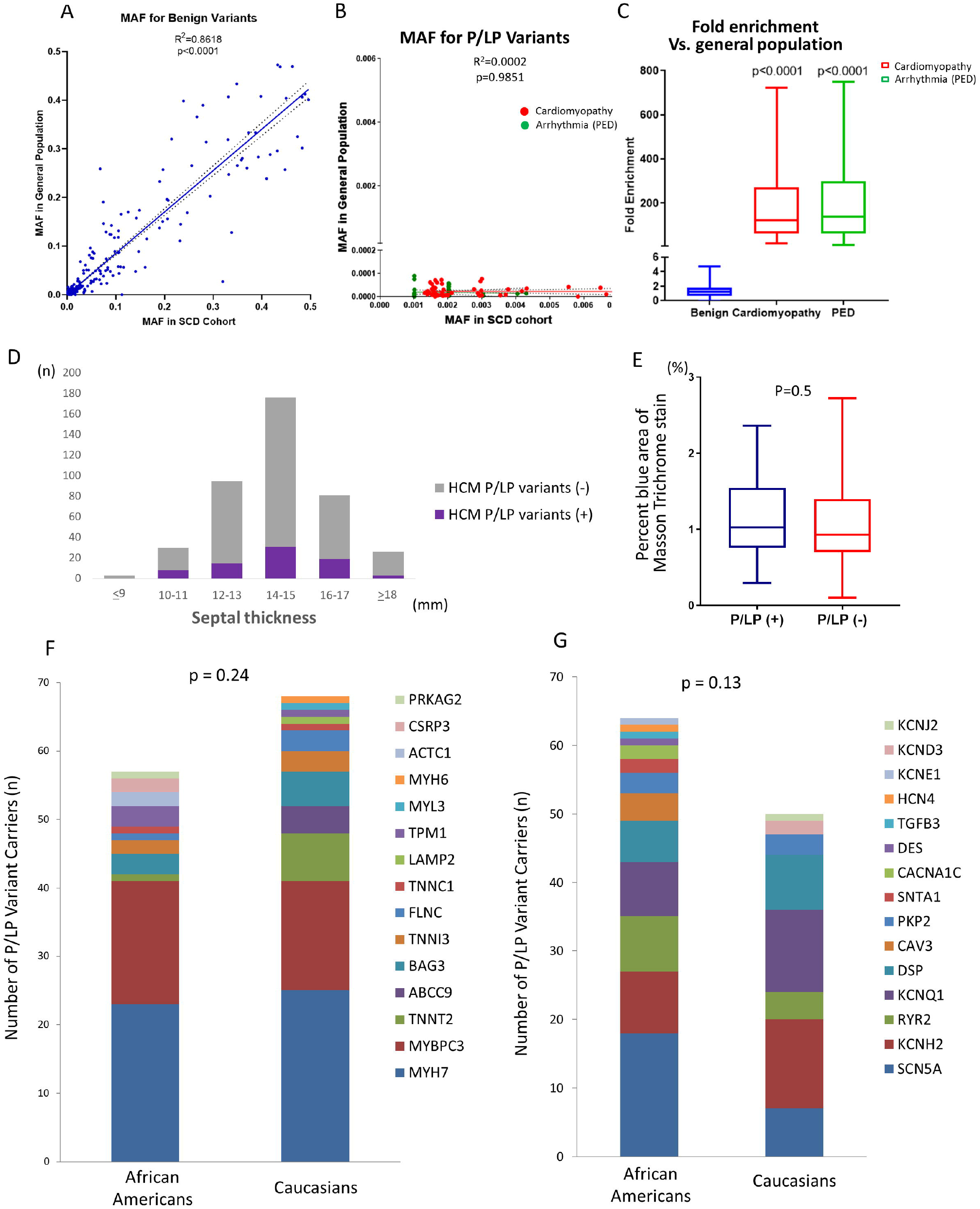
Racial Disparities in Cardiomyopathy and Arrhythmia Gene Variants. **A-B**. Comparison of minor allele frequency (MAF) for benign and P/LP variants between unexplained SCD cohort and general population using Spearman’s correlation. Note the MAF for benign variants was linear in nature (A) whereas the MAF for P/LP was much more common in our unexplained-SCD cohort versus the general population (B). **C**. MAF for P/LP showed more than 121.0 and 138.5 -fold enrichment vs. general population in for cardiomyopathy and arrhythmia P/LP variants whereas benign variants were not enriched (1.2). **D**. Distribution graph of HCM P/LP variants stratified by septal thickness (i.e. 2 mm increments) demonstrated equal distribution of variants. **E**. The percent area myocardial fibrosis in subjects with and without P/LP HCM gene variants was similar. **F-G**. Numbers of the carriers for each P/LP variants per gene in African Americans versus Caucasians in cardiomyopathy genes (D) and arrhythmia genes (E).

### Clinical Characteristics and Heart Dimensions in Subjects With or Without Pathogenic /Likely Pathogenic Gene Variants

No significant difference was found in clinical characteristics and heart dimensions between subjects with or without P/LP variants of either male or female sex (Table 1). Overall, among subjects carrying P/LP variants, 48% were African Americans and 52% Caucasians, 40% were female and 60% male.

**Table 1.** Patient Characteristics in Subjects with and Without Pathogenic / Likely Pathogenic (P/LP) Gene Variants.

| Patient Characteristics | P/LP variant (+) n=143 |  | P/LP variant (-) n=271 |  | p value |  |
| --- | --- | --- | --- | --- | --- | --- |
| Age | 39.4 (28.0-48.0) |  | 41.2 (31.0-48.0) |  | 0.22 |  |
| Sex (Female), n (%) | 57 (39.9%) |  | 98 (36.2%) |  | 0.52 |  |
| Race (African American), n (%) | 68 (47.6%) |  | 140 (51.7%) |  | 0.47 |  |
| Body / Heart Dimensions | Women n=57 | Men n=86 | Women n=98 | Men n=173 | P<br>(women) | P<br>(men) |
| Body height, cm | 165.1 (162.6-172.7) | 177.8 (171.5-180.3) | 167.6 (162.6-172.1) | 177.8 (172.7-184.8) | 0.82 | 0.37 |
| Body Weight, kg | 75.8 (63.5-100.0) | 83.5 (69.9-94.1) | 80.3 (62.6-96.6) | 83.0 (71.2-97.8) | 0.96 | 0.49 |
| Body Mass Index, kg/m<sup>2</sup> | 27.3 (22.7-35.9) | 27.0 (22.6-29.2) | 29.0 (23.3-34.6) | 26.5 (22.6-30.2) | 0.98 | 0.70 |
| Body Surface Area, m<sup>2</sup> | 1.75 (1.57-2.03) | 1.89 (1.68-2.02) | 1.80 (1.60-1.96) | 1.90 (1.73-2.07) | 0.87 | 0.48 |
| Heart Weight, g | 360.0 (302.5-425.0) | 405.0 (375.0-450) | 350.0 (300.0-420.0) | 430.0 (380.0-466) | 0.81 | 0.10 |
| LV diameter, mm | 35.0 (30.0-40.0) | 32 (29-40) | 35.0 (28.5-40.0) | 35.0 (30.0-40.0) | 0.99 | 0.39 |
| LV free wall thickness, mm | 13.0 (11.0-15.0) | 13.0 (12.0-15.0) | 12.0 (10.0-14.0) | 14.0 (12.0-15.0) | 0.06 | 0.18 |
| Septum thickness, mm | 14.0 (12.0-15.0) | 15.0 (14.0-16.0) | 14.0 (12.0-15.0) | 15.0 (14.0-17.0) | 0.26 | 0.73 |
| RV thickness, mm | 4.0 (4.0-5.0) | 4.0 (4.0-5.0) | 4.0 (4.0-5.0) | 5.0 (4.0-5.0) | 0.56 | 0.30 |
| Septum / Free wall ratio | 1.08 (1.00-1.16) | 1.11 (1.06-1.18) | 1.13 (1.10-1.20) | 1.08 (1.00-1.19) | 0.16 | 0.19 |

LV=left ventricle, RV=right ventricle, BW=body weight. Data are shown at median (IQR) unless indicated.

Because of the high overall frequency of subjects with unexplained SCD carrying HCM P/LP variants (i.e. 79/413, 19.1%), we conducted further sub-analysis in this group. We compared heart/body dimensions in study cohort and subjects with HCM P/LP variants to 49 hearts with pathologic diagnosis of HCM (from our collection), and found significant differences in most heart measurements (Supplemental Table 4). When we examined the distribution of HCM P/LP variants stratified by septal thickness across all cases of unexplained SCD, there was equal distribution of HCM P/LP across all septal thicknesses (Figure 3D). Sections of left ventricular myocardium from 21 consecutive subjects with P/LP HCM gene variants and 22 consecutive subjects without P/LP gene variants (median age; 39 vs. 31, respectively, p=0.2) were evaluated for evidence of myocardial fibrosis. The percent area myocardial fibrosis in subjects with and without P/LP HCM gene variants was similar [1.02 (0.76-1.55) % vs. 0.93 (0.70-1.40) %, respectively, p=0.5, Figure 3E, Supplemental Table 5]. Thus, in subjects with unexplained SCD, there was no relationship between heart dimensions, myocardial fibrosis and the presence of P/LP HCM variants.

### Racial Differences

Similar numbers of African Americans carried P/LP gene variants versus Caucasians (68/208 [32.7%] vs. 75/205 [36.6%], respectively, p = 0.5) (Table 1). The numbers of P/LP gene variant carriers by race are shown for both arrhythmia and cardiomyopathy genes in Figure 3F and G, respectively. The most frequent P/LP variants in cardiomyopathy genes were MYBPC3 and MYH7, similar between two races; however, the most frequent P/LP variants in arrhythmia genes were different-SCN5A in African Americans and KCNH2/KCNQ1 in Caucasians (Figure 3 F-G). Most of the P/LP variants were exclusive to one race [African Americans 41/75 (54.7%), Caucasians 27/75 (36.0%) in arrhythmia genes; African Americans 33/84 (39.3%), Caucasians 39/84 (46.4%) in cardiomyopathy genes], whereas only 7/84 (9.3%) and 12/84 (14.3%) of P/LP variants were seen in both races for arrhythmia and cardiomyopathy genes, respectively (Supplemental Figure 2A-B).

MAF for benign variants in African American versus Caucasians was linear (R^2^=0.6607, p<0.0001) (Supplemental Figure 2C). However, MAF for variants with unknown significance (VUS) were commonly seen in African Americans (R^2^=0.0018, p<0.12) (Supplemental Figure 2D). We identified 3 VUS (2 in MYH7 and 1 in JUP) only found in African Americans, which were >100-fold enriched in our cohort compared to the general population, suggesting they could be associated with damaging function and risk of unexplained-SCD (Table 2).

**Table 2.**
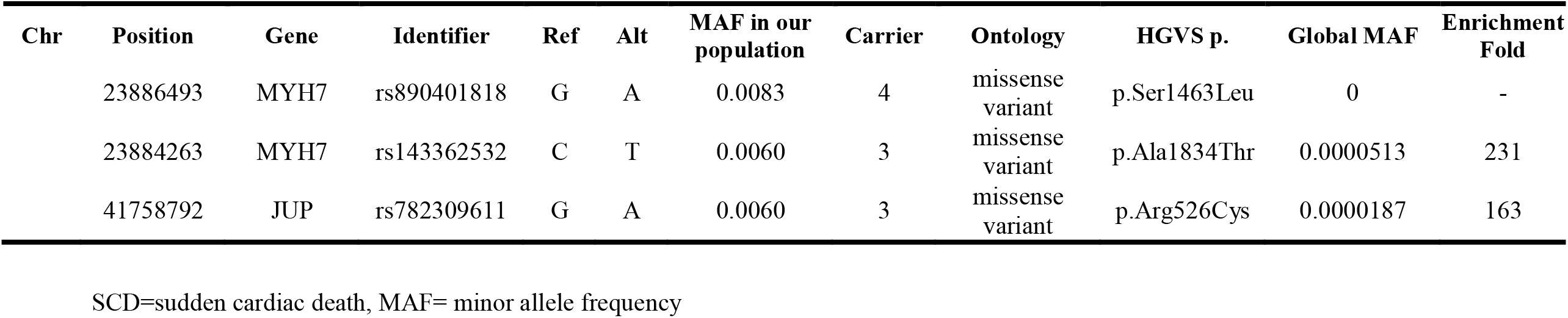
Variants with Unknown Significance in African American only with >100X enriched in SCD Cohort.

## DISCUSSION

SCD remains a major public health problem worldwide and is estimated to account for 15-20% of all deaths ^1, 2^. The current study examined the association between cardiomyopathy and arrhythmia genetic variants and unexplained-SCD and looked at whether race-based differences in these genetic variants may exist. To the best of our knowledge this is the largest and most comprehensive genetic autopsy study ever to be performed in the US. In our autopsy registry of over 5,000 cases of suspected sudden cardiac death referred from OCME-MD, we identified 683 hearts from Caucasians and African Americans with unexplained SCD. Genetic analysis of 29 cardiomyopathy and 39 primary arrhythmia gene variants was performed in 413 subjects and showed that slightly greater than one-third of subjects dying of unexplained-SCD in the State of Maryland carried P/LP variants for cardiomyopathy or arrhythmia genes, representing a greater than 100-fold enrichment in the frequency of these variants versus the general population.

Although our results were not combined with screening of surviving family members, our data suggests that genetic causes may contribute to a far greater number of unexplained sudden cardiac death cases in adults than previously appreciated. Although the relationship between PED gene variants and risk of unexplained SCD is well recognized especially in younger adults ^20-22^, one surprising finding of our study was the large numbers of subjects with unexplained SCD carrying genetic variants for HCM (19.1%), which suggests a potential association of these genes with risk of SCD in the absence of structural heart disease. In addition, our study is the first to examine the frequency of P/LP variants in victims of unexplained SCD of African American and Caucasian ancestry. Overall, African Americans had a similar frequency of P/LP HCM gene variants versus Caucasians, but most variants were exclusive by race and African Americans were more likely to harbor variants of unknown significance.

Previous autopsy studies of victims of sudden cardiac death have shown significant variation in the numbers of subjects without abnormal cardiac exam sometimes as low as 5% when older subjects are included. In one series with subjects similar in age (mean age 38) to our cohort, 187 of 902 cases (21%) had no evidence of cardiac pathology ^23^. We report a slightly lower percent of patients with unexplained SCD (13%) although we excluded from evaluation younger (i.e. <18 years of age), non-Caucasian, and non-African American subjects.

Autopsy studies have demonstrated the utility of performing genetic analysis of subjects with unexplained-SCD, yielding diagnoses in up to one-third of young SCD cases. In the largest series of 302 cases with unexplained sudden cardiac death (median age 24 years), molecular autopsy for 77 genes for PED and cardiomyopathies, a P/LP variant was identified in 40 cases (13%). The most common variants found were in gene for PED (catecholaminergic polymorphic ventricular tachycardia (6%)) and congenital long QT syndrome (4%) with only 2% harboring P/LP cardiomyopathy gene variants^22^. These data were generated from a multiple sources including routine autopsy referrals from multiple medical centers in the Europe (United Kingdom, New Zealand, Denmark, and Netherlands) which might affect diagnostic yield of molecular autopsy since these were not all suspected sudden cardiac deaths. Another study of sudden cardiac death which was prospective in nature consisted of autopsies from all cases of sudden cardiac death in children and young adults ages 1 to 35 years in Australia and New Zealand. In cases of unexplained-SCD, a clinically relevant gene mutation was found in 27% of cases with 17/131 carrying PED variants, 19/113 carrying cardiomyopathy variants, and 2/113 carrying both ^7^. Although the definition of P/LP variants was different in this versus the present study, both demonstrated the well-known association between PED and unexplained SCD. However, both also report the same unexpected finding that a significant number of subjects dying of unexplained SCD harbored cardiomyopathy variants.

Some may question whether carriers of P/LP variants for cardiomyopathy genes in this study harbored underlying structural heart disease that might have been missed on autopsy. In the current study, myocardial sections from at least 6 parts from the heart were routinely taken in all cases, and a detailed histological examination was performed to rule out presence of any cardiomyopathic process. Maron et al. demonstrated that ventricular septal and free wall thickness were thicker in autopsy hearts when the thickness was compared with diastolic thickness measured by echocardiogram that was performed prior to death ^16^. Therefore, wall thickness criteria used in clinical echocardiography is not used as a diagnostic criteria for cardiomyopathies during autopsy examination. Instead, increased septal/free wall ratio (≥1.3) and/or abnormal histopathologic findings such as myofiber disarray, myocyte hypertrophy, fibrosis (scarring), and thickening of intramural coronary arteries are routinely used ^24-29^. Moreover, wall thickness measurements in our study were similar to what was shown in a previous study evaluated cardiac morphology of 765 normal hearts ^15^. Heart weight, which is known to become heavier as body mass index (BMI) or body weight increases ^15, 30^, was also consistent with large autopsy studies of normal hearts ^15, 31^.

Even in the present day, SCD risk prediction remains difficult without precise factors known to increase risk of death on an individual basis, especially in those without overt cardiac disease. Yet, relatively few studies have been conducted in part because of the difficulty of performing a full cardiac autopsy on a consistent basis and in a large number of subjects. Our data in this study from the CVPath Sudden Death Registry represents the largest unexplained-SCD study performed to date. Of the cardiomyopathy genes studied, the majority of mutations detected were considered P/LP for HCM. Although it is a common genetic heart disorder and well-recognized cause of SCD in those with overt heart disease, the role of genetics in risk prediction remains uncertain, in those without clinically detectable disease ^4, 5^. The prevailing clinical perception is that sudden death risk in HCM is related to the presence of ventricular hypertrophy. However, case reports have documented the occurrence of SCD in those carrying genetic HCM variants but without overt clinical disease ^32^. There is currently disagreement by consensus expert panels about the significance HCM P/LP variants in those without overt cardiac disease ^4, 5^.

Studies of single families with specific variants are limited by numbers, length of follow-up, and the possibility that shared genetic and yet unknown environmental modifiers may affect overall risk. Indeed, large scale studies regarding the rate of SCD in individuals with genotype positive phenotype negative have not been performed. The concordance in findings of the association of cardiomyopathy P/LP variants and unexplained SCD between the study of Bagnall and our own, the two largest studies to specifically examine the genetics of unexplained SCD, suggests further work is needed to understand whether carrying certain cardiomyopathy genes without overt clinical disease increases risk for unexplained SCD.

Study of different and underrepresented races (i.e. African Americans) is another unique aspect of our study. Overall, African Americans harbored a similar frequency of P/LP variants versus Caucasians for both cardiomyopathies and PED. While the most frequent P/LP variants in cardiomyopathy genes were MYBPC3 and MYH7 for both races; most frequent P/LP variants in arrhythmia genes were different-SCN5A in African Americans and KCNH2/KCNQ1 in Caucasians with most of the P/LP variants were exclusive to one race. (Only 9.3% and 14.3% of P/LP variants were seen in both races for arrhythmia and cardiomyopathy genes, respectively.) Moreover, the prevalence in variants of uncertain significance were higher in African Americans compared with Caucasians, which is consistent with the previous study ^33^. In our study, we found in total 3 VUS which were >100x enriched in our African American unexplained-SCD cohort compared to general population, suggesting a causal role in risk for unexplained SCD.

Our study has several limitations. First, a detailed family history of SCD was lacking and thus the data collected relied only on the association of the presence of a variant and the occurrence of death in one individual rather than within a familial cohort. Second, while all the cases of the current study were collected from OCME-MD, there may be a referral bias and thus the study findings may not accurately represent the frequency of genetic variants in victims of SCD in the community.

In conclusion, this study represents one of the largest examinations reported of the association between causative cardiomyopathy and arrhythmia genetic variants and unexplained-SCD. Over one-third of those with unexplained-SCD were carriers of P/LP variants. Our findings with respect to both the association of unexplained SCD with cardiomyopathy genes and race-specific genetic variants suggest new avenues of study for this poorly understood entity.

## Data Availability

All data referred to in the manuscript are available.

## Abbreviations

SD: Sudden Death
SCD: Sudden Cardiac Death
P/LP: Pathogenic or Likely Pathogenic
CoD: Cause of Death
HCM: Hypertrophic Cardiomyopathy
DCM: Dilated Cardiomyopathy
LQTS: Long QT Syndrome
ARVD: Arrhythmogenic Right Ventricular Dysplasia

## Author Contributions

LG, ST, AVF contributed to the design of the study. RF, REB contributed to the sequencing library preparation. LG, ST, KHP, CMM, RZ, DTF, CCH, AVF contributed to data analysis. RK, MER, RV contributed to cardiac pathologic assessment. LG, ST, PJL, LW, contributed to the analysis of myocardial fibrosis. MDK, CJC, NG, BA, DRA, MB contributed to DNA extraction. HJ, AS, YS, HM, AC, SK, JP, LW, SNH, AG, FDK, MER, BDM, DS, DRF, CCH, participated in scientific discussions. LG, ST, RV, AVF drafted and/or edited the manuscript, and was critically reviewed by all the authors.

## Acknowledgements

This study is supported by the CVPath Institute Research Fund.

---

**Supplemental Figure 1.**
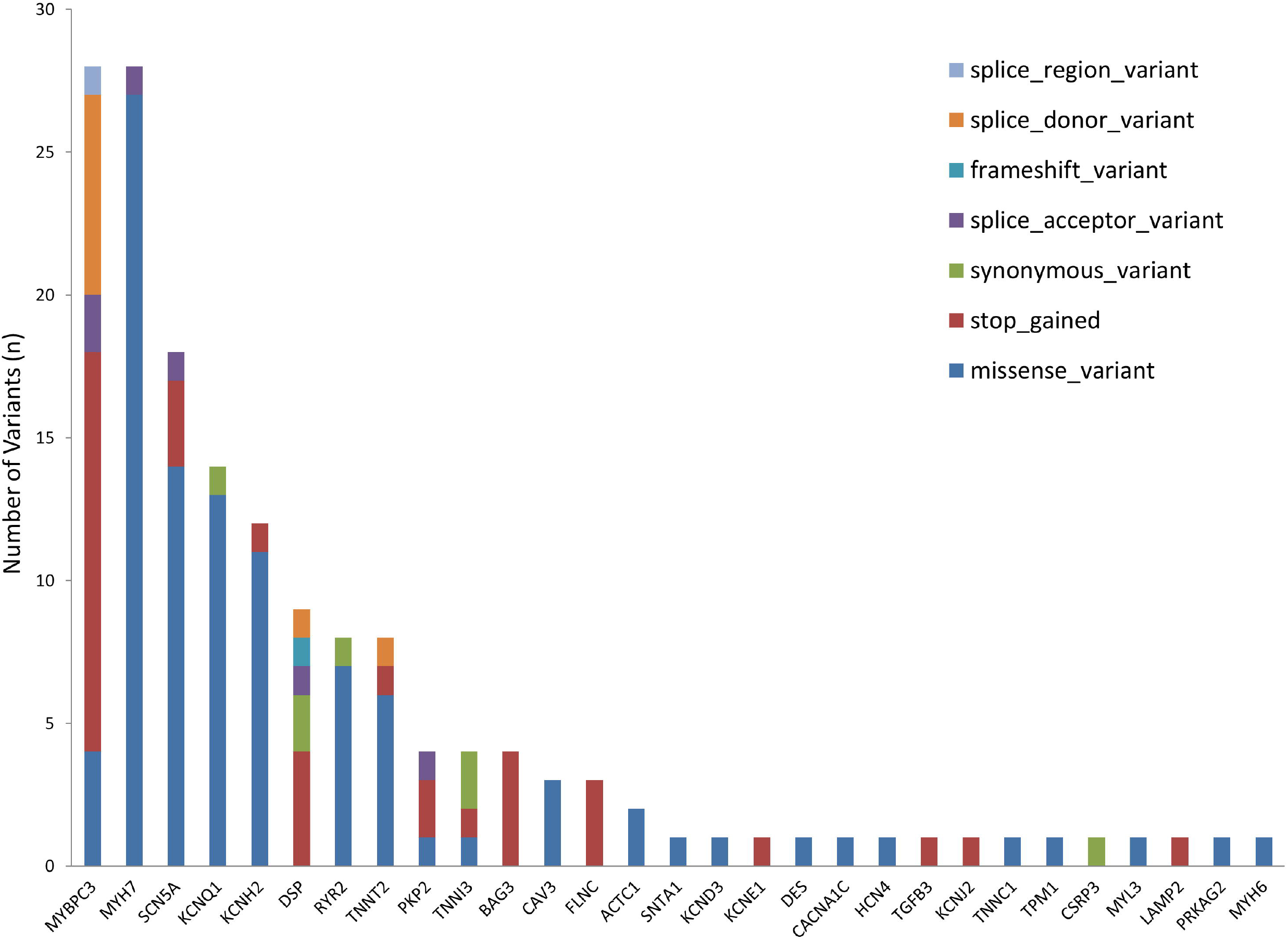

**Supplemental Figure 2.**
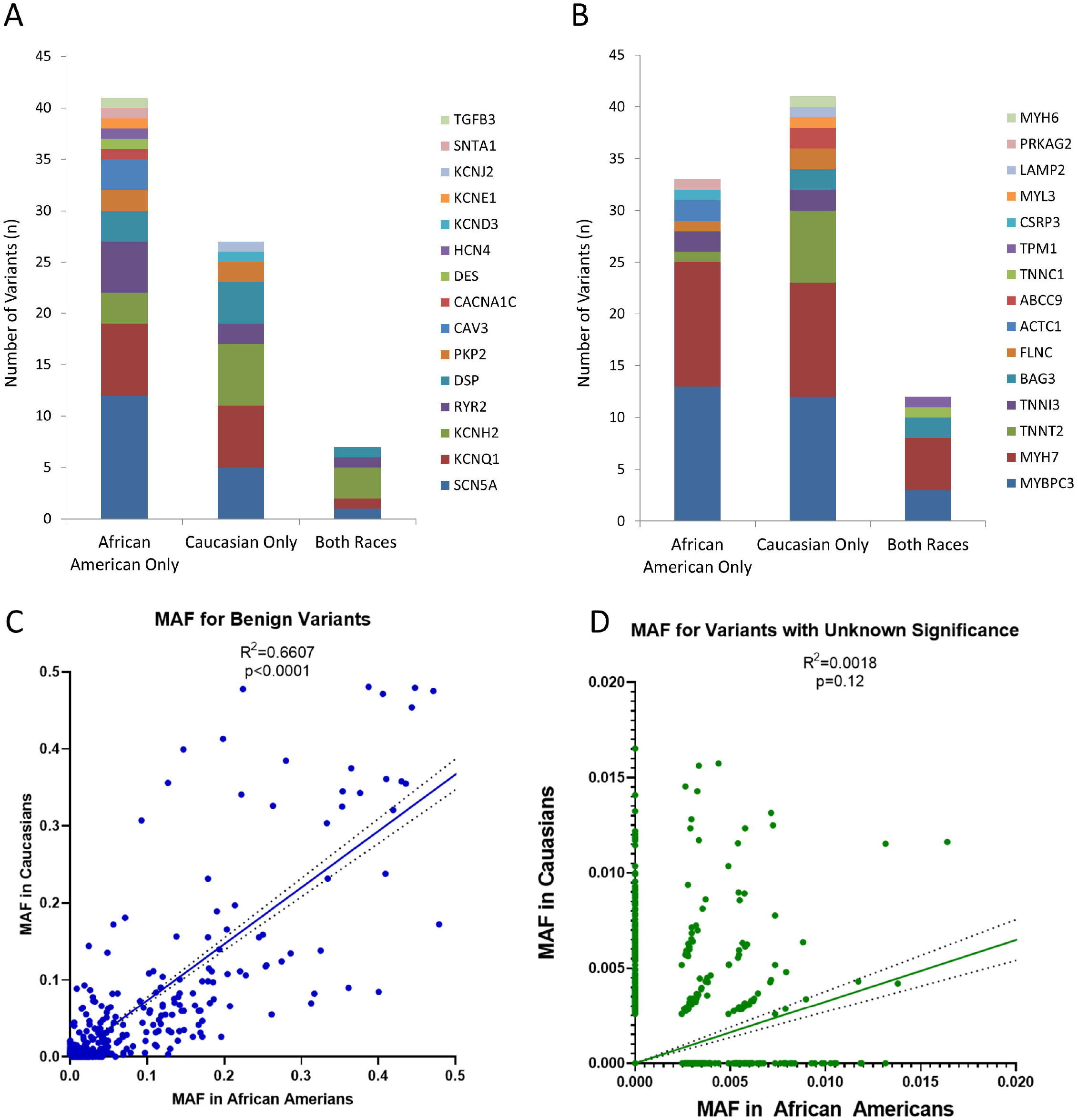

